# Whole-genome analysis of Malawian *Plasmodium falciparum* isolates identifies potential targets of allele-specific immunity to clinical malaria

**DOI:** 10.1101/2020.09.16.20196253

**Authors:** Zalak Shah, Myo T. Naung, Kara A. Moser, Matthew Adams, Andrea G. Buchwald, Ankit Dwivedi, Amed Ouattara, Karl B Seydel, Don P. Mathanga, Alyssa E. Barry, David Serre, Miriam K. Laufer, Joana C. Silva, Shannon Takala-Harrison

## Abstract

Individuals acquire immunity to clinical malaria after repeated *Plasmodium falciparum* infections. This immunity to disease is thought to reflect the acquisition of a repertoire of responses to multiple alleles in diverse parasite antigens. In previous studies, we identified polymorphic sites within individual antigens that are associated with parasite immune evasion by examining antigen allele dynamics in individuals followed longitudinally. Here we expand this approach by analyzing genome-wide polymorphisms using whole genome sequence data from 140 parasite isolates representing malaria cases from a longitudinal study in Malawi and identify 25 genes that encode likely targets of naturally acquired immunity and that should be further characterized for their potential as vaccine candidates.

## INTRODUCTION

Despite recent progress in reducing the burden of malaria, this disease remains a leading cause of mortality worldwide, resulting in an estimated 405,000 deaths in 2018 (*WHO* | *World Malaria Report 2018*, n.d.). In areas with high transmission of *Plasmodium falciparum*, individuals develop immunity to malaria (Marsh & Kinyanjui, 2006). This immunity does not provide sterile protection against all infections, but decreases the risk of clinical disease, and increases with age as individuals are repeatedly exposed to the parasite (Cowman et al., 2016; Ryg-Cornejo et al., 2016). This age-related pattern of immunity to disease is thought to reflect the need for a repertoire of immune responses to multiple alleles in diverse parasite antigens (Buchwald, Sorkin, et al., 2019; Cowman et al., 2016; Marsh & Kinyanjui, 2006; Portugal et al., 2013). The extensive genetic diversity in *P. falciparum* surface antigens is thought to have evolved over millennia as a means of parasite immune evasion (Weedall & Conway, 2010). Allele-specific immune responses have been demonstrated for several parasite antigens (Barry et al., 2011; Cortés et al., 2005; Crompton et al., 2010; Dutta et al., 2007; Early et al., 2018; Osier et al., 2008; Polley et al., 2007; Tran et al., 2014). In previous work, we examined parasite alleles in repeated infections occurring in individuals followed longitudinally and identified specific polymorphic sites within parasite surface antigens (i.e. AMA1 and MSP1) where amino acid changes were associated with immune escape and increased risk of disease, consistent with allele-specific acquisition of immunity to these antigens (Takala et al., 2007, 2009). Furthermore, malaria subunit vaccines based on a single antigen allele have displayed greater efficacy against parasites with alleles matching the vaccine strain compared to the diverse alleles observed in natural parasite populations (Ouattara et al., 2013, 2015; Takala et al., 2009; Thera et al., 2011). Such allele-specific vaccine efficacy could lead to poor overall vaccine efficacy when the vaccine target allele is at low frequency in the parasite population and could result in selection of non-vaccine alleles capable of vaccine escape (Ouattara et al., 2015). Overcoming this scenario may require the design of a multivalent malaria vaccine (Ouattara et al., 2015). However, the design of such a vaccine is hampered by an incomplete knowledge of which parasite proteins are targets of acquired natural immunity.

The advent of technologies allowing whole genome sequencing at epidemiological scales has allowed investigators to transition from investigation of single antigens to performing genome-wide screens to identify loci likely to be involved in the acquisition of protective immunity to malaria, including uncharacterized genes encoding products of unknown function (Gardner et al., 2002; *PlasmoDB : The Plasmodium Genomics Resource*, n.d.). Although there have been previous genome-wide studies in *P. falciparum* to identify genomic signatures of balancing or diversifying selection at a population level (Amambua-Ngwa et al., 2012; Mobegi et al., 2014; Mu et al., 2007), these studies do not associate identified signatures with clinical outcomes at an individual level, making it difficult to directly link such signatures to immune selection. Expanding on our previous approach where we examined the dynamics of vaccine antigen alleles in individuals’ repeated infections over time in relation to the development of symptoms (Takala et al., 2007, 2009), we compared whole genome sequence data generated from *P. falciparum* infections collected from participants in a longitudinal cohort study conducted in Malawi to identify targets of allele-specific immunity to malaria. Specifically, we compared the frequency of parasite alleles in symptomatic infections occurring in individuals with different levels of malaria immunity to identify significantly differentiated sites, and also compared alleles in repeated infections within an individual *versus* between individuals to identify polymorphic sites that vary most within individuals. Genes identified using both approaches were considered likely immune targets and were further examined for their potential as vaccine candidates. As a proof of concept of the utility of our approach in identifying targets of allele-specific immune responses, we compared the frequency of alleles in one of the identified antigens (previously considered as a potential vaccine candidate) in individuals with different levels of malaria immunity to test the hypothesis that individuals who are more immune become ill when infected with a parasite having rarer alleles to which they have not yet developed immunity.

## RESULTS

### Participant/infection characteristics and definition of immune status

To identify targets of allele-specific immunity to malaria, we generated whole-genome sequence data from 140 parasite isolates collected from symptomatic infections occurring in participants in a longitudinal cohort study in Malawi (Buchwald, Sixpence, et al., 2019).

Although age is often used as a proxy for immune status in high transmission areas (Egan et al., 1996; Metzger et al., 2003; Perraut et al., 2017; Tran et al., 2014), this metric does not account for heterogeneous exposure to infectious mosquito bites, which has been observed in endemic areas (Bejon et al., 2010; Bousema et al., 2010; Clark et al., 2008; Elissa et al., 2003; Gaudart et al., 2006; Kang et al., 2018). To better account for heterogeneous exposure at an individual level, we used the proportion of total infections that were symptomatic over the two-year study period to categorize immune status, using the median as a cutoff to define high and low immunity groups. Although these groups consisted of individuals with a range of ages, the median age (13.18 years) of individuals in the group with high immunity was significantly greater than the median age (7.27 years) of individuals in the low immunity group (*p-*value =0.0001, Wilcoxon rank sum test) (Figure 1, Table 1), as would be expected in a high malaria transmission setting such as Malawi.

**Table 1.** Participant/infection characteristics in high and low immunity groups^#^.

| Characteristics | High Immunity<br>($n=28$) | Low Immunity<br>($n=33$) | $P$-value |
| --- | --- | --- | --- |
| Median age (years) | 13.18 | 7.27 | 0.0002<sup>+</sup> |
| Male (%) | 36 | 52 | 0.33<sup>*</sup> |
| Median parasitemia (parasites/uL) | 4710 | 48200 | 7.74e-06<sup>+</sup> |
| Median genome coverage 20x (%) | 88.73 | 90.14 | 0.2673<sup>+</sup> |
| Median depth of coverage | 130x | 156x | 0.2485<sup>+</sup> |

<sup>#</sup>High complexity infections lacking a predominant parasite clone were excluded
<sup>\*</sup> $P$ -value determined using z-score test for difference in proportions
<sup>+</sup> $P$ -value determined using Wilcoxon rank sum test

**Figure 1.**
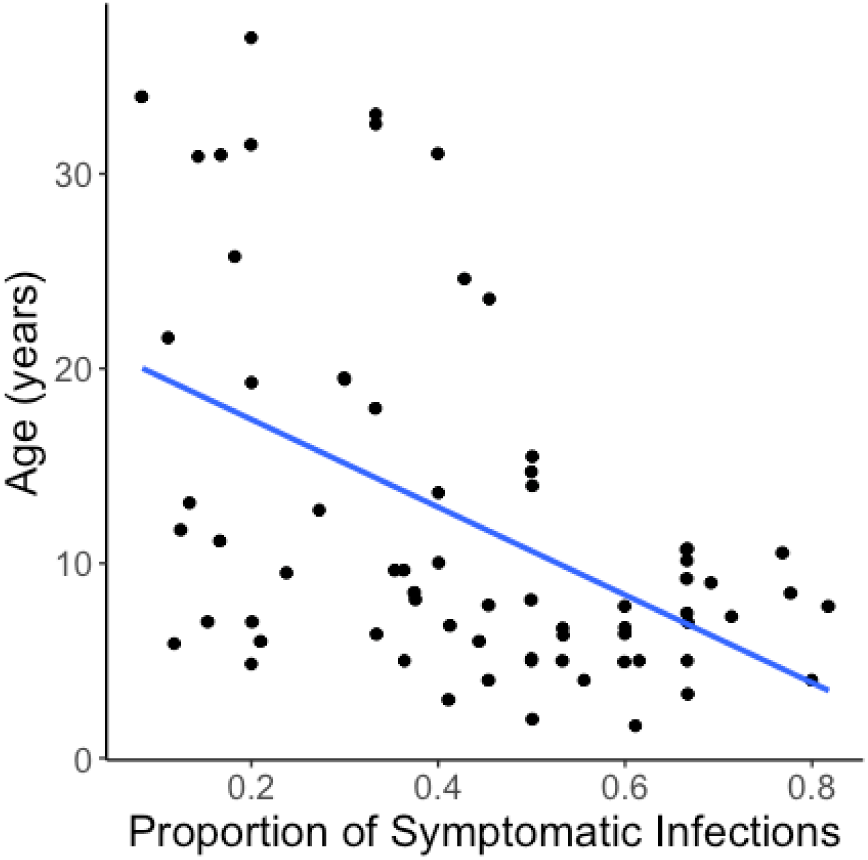
Relationship between proportion of symptomatic infections and age. Scatterplot, including linear regression line (blue), shows the relationship between the proportion of symptomatic infections per individual over the course of the study and age of the individual at enrollment.

Only one infection from each individual was included in comparisons between the high and low immunity groups, with samples selected in a manner to reduce temporal variability between infections (see Methods). DEploid-IBD (Zhu et al., 2017) was used to estimate the proportion of each clone within an infection. Infections without a predominant clone (i.e., where the majority clone had a frequency <60% within the infection) were defined as complex infections. Although the median frequency of the majority clone was not significantly different between infections in the two immunity groups (Supplementary Fig 1, *p-*value = 0.3372, Wilcoxon rank sum test), the high immunity group had a greater number of complex infections (n=7) than the low immunity group (n=2). These nine complex infections were excluded from further analysis to avoid confounding by infection complexity and misclassification of alleles likely contributing to clinical illness.

The median parasite density of infections in the high immunity group was significantly lower than in the low immunity group (Table 1, *p-*value = 7.737 × 10^−06^, Wilcoxon rank sum test). However, there was no significant difference in the percentage of the parasite genome with at least 20-fold coverage (Table 1, *p-*value = 0.2673, Wilcoxon rank sum test), or in the median average depth of coverage (Table 1, *p-*value = 0.2485, Wilcoxon rank sum test) between whole genome sequence data generated from infections in the two groups.

### Differentiated loci between groups with different levels of immunity to clinical malaria

We hypothesized that, because of allele-specific immune responses, individuals with greater protective immunity to malaria would experience disease when infected with parasite antigen alleles that are rarer in the parasite population, having already developed immunity to more common alleles circulating in the population. Thus, we would expect significant genetic differentiation between antigen alleles in individuals with high *versus* low immunity at loci that are targets of allele-specific immunity.

To test this hypothesis, Wright’s fixation index (*F*_ST_), a measure of genetic differentiation between two populations (Bhatia et al., 2013), was estimated per non-synonymous single nucleotide polymorphism (SNP) to identify genetically differentiated sites between parasites from the high and low immunity groups with the significance threshold based on 10,000 permutations. We identified 160 sites (in 145 genes) in the parasite genome that were significantly differentiated between the two immunity groups (*p-*value ≤ 0.0095, Figure 2a, Supplementary Table 1). Fifty-five of the genes containing significantly differentiated sites (38%) encode proteins of unknown function that are not associated with any computed or curated molecular function or biological process based on Gene Ontology (Supplementary Table 1). These 145 gene products included some proteins previously identified as potential vaccine candidates, including AMA1 (apical membrane antigen 1) (Takala et al., 2009; Thera et al., 2011, p. 20), ASP (apical sushi protein, PF3D7_0405900) (Vanegas et al., 2014), CLAG8 (cytoadherence linked asexual protein 8, PF3D7_0831600) (Gupta et al., 2015; Iriko et al., 2008), SLARP (sporozoite and liver asparagine-rich protein, PF3D7_1147000) (van Schaijk et al., 2014) and a conserved protein of unknown function (PF3D7_1359000) (Krzyczmonik et al., 2012).

**Figure 2.**
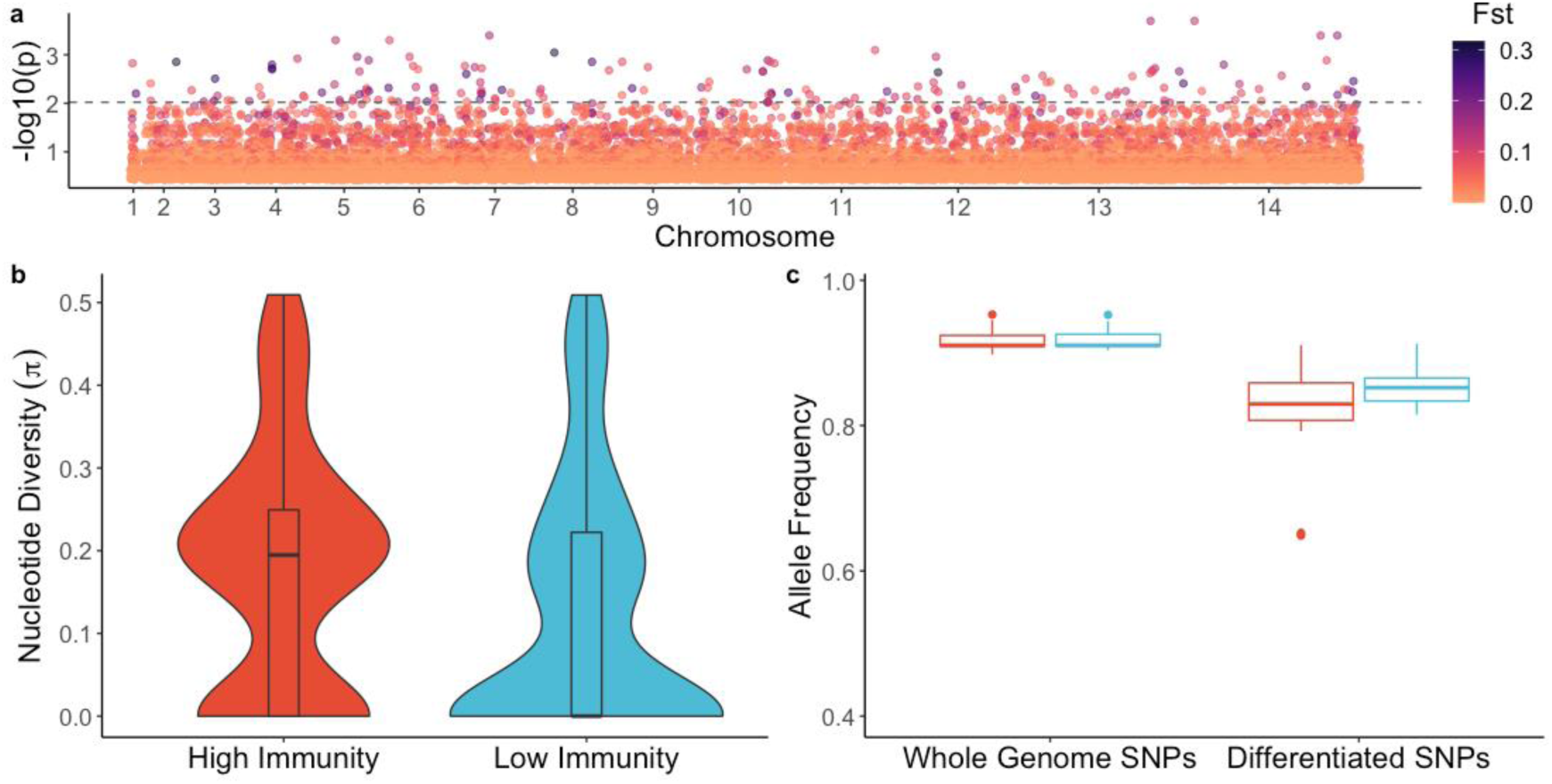
Genetic differentiation between parasites from high immunity *vs*. low immunity groups. a) Genome-wide genetic differentiation (*F*_ST_) between parasites from individuals with higher immunity *vs*. lower immunity. Each point represents a variable, non-synonymous site. Results are plotted as –log_10_ *p*-values on the y-axis. The color of each point represents the *F*_ST_ value, with darker points indicating higher *F*_ST_ values. The dashed line denotes statistical significance (*p-*value = 0.0095), with *p*-value determined by permutation. b) Nucleotide diversity for significantly differentiated SNPs in parasites from individuals with higher immunity and lower immunity. c) Box-plot of mean allele frequency per individual based on SNPs in the whole genome sequences, and which are significantly differentiated SNPs from (a). Red indicates the high immunity group and blue color indicates the low immunity group.

To further test the hypothesis that individuals who are more immune become symptomatic when infected with parasites having antigen alleles that are rarer in the parasite population, we estimated nucleotide diversity at significantly differentiated non-synonymous sites in parasites from the two immunity groups and observed a significantly greater median nucleotide diversity in parasites from the high immunity group compared to parasites from the low immunity group (Figure 2b, *p-*value= 4.74 ⨯ 10^−05^, Wilcoxon rank sum test). In addition, we estimated the average frequency of alleles in each infection at both significantly differentiated sites and genome-wide variable sites. The median frequency of alleles at genome-wide variable sites was not significantly different between immunity groups (Figure 2c, *p-*value = 0.8801, Wilcoxon rank sum test); in contrast, at the differentiated sites, the median frequency of alleles was significantly lower in the high immunity group compared to the low immunity group (Figure 2c, *p-*value = 0.0072, Wilcoxon rank sum test). These results are consistent with the scenario that individuals in the high immunity group are infected with parasites having different lower-frequency alleles compared to individuals in the low immunity group who are infected with parasites sharing more common alleles.

Within 23 polyclonal infections, we also compared the proportion of mismatched alleles between the predominant and minor clones at both significantly differentiated sites and genome-wide variable sites in order to assess whether these clones differ at sites thought to be relevant for immunity. We observed a significantly greater median proportion of mismatches between the major and minor clones within an infection at the differentiated sites compared to genome-wide variable sites (Supplementary Figure 2, *p-*value = 8 × 10^−05^, Wilcoxon rank sum test), consistent with the hypothesis that the predominant clone represents a breakthrough infection that has escaped allele-specific immune responses that maintain minor clones at a subclinical level.

### Loci that differ more within individuals than between individuals

We expected that allele-specific immune responses would result in a greater proportion of genetic differences in parasites causing multiple symptomatic infections within an individual compared to parasites causing infection in different individuals, at antigenic loci that are targets of immunity. To identify regions of the parasite genome that are most different in infections occurring within an individual *versus* between individuals, we compared the allelic states at each variable non-synonymous site across the genome between pairs of isolates sampled within an individual and between individuals and estimated the proportion of mismatches per site in each group (Figure 3a). The distribution of the difference in the proportion of mismatches across all non-synonymous variable sites between the two groups (within minus between) is shown in Figure 3b (see also Supplementary Fig 3a). The difference in the proportion of mismatches at each site had a median and mode equal to zero, indicating that the proportion of mismatches was not different within and between individuals at most sites. There was no significant correlation between the number of days between infections in a pair and the proportion of mismatches (Supplementary Fig 2b, Pearson’s correlation *r* = 0.065, *p*-value = 0.3043), suggesting that time was not a significant confounding factor in the analysis. We further examined the top 1% of sites that differed most within individuals compared to between individuals, which included 223 SNPs, located in 173 genes (Supplementary Table 2). Sixty-eight (39%) of these 173 genes encode proteins of unknown function that are not associated with any computed or curated molecular function or biological process based on Gene Ontology, and at least 15 (8.7%) encode for proteins that have been previously identified as potential vaccine candidates.

**Figure 3.**
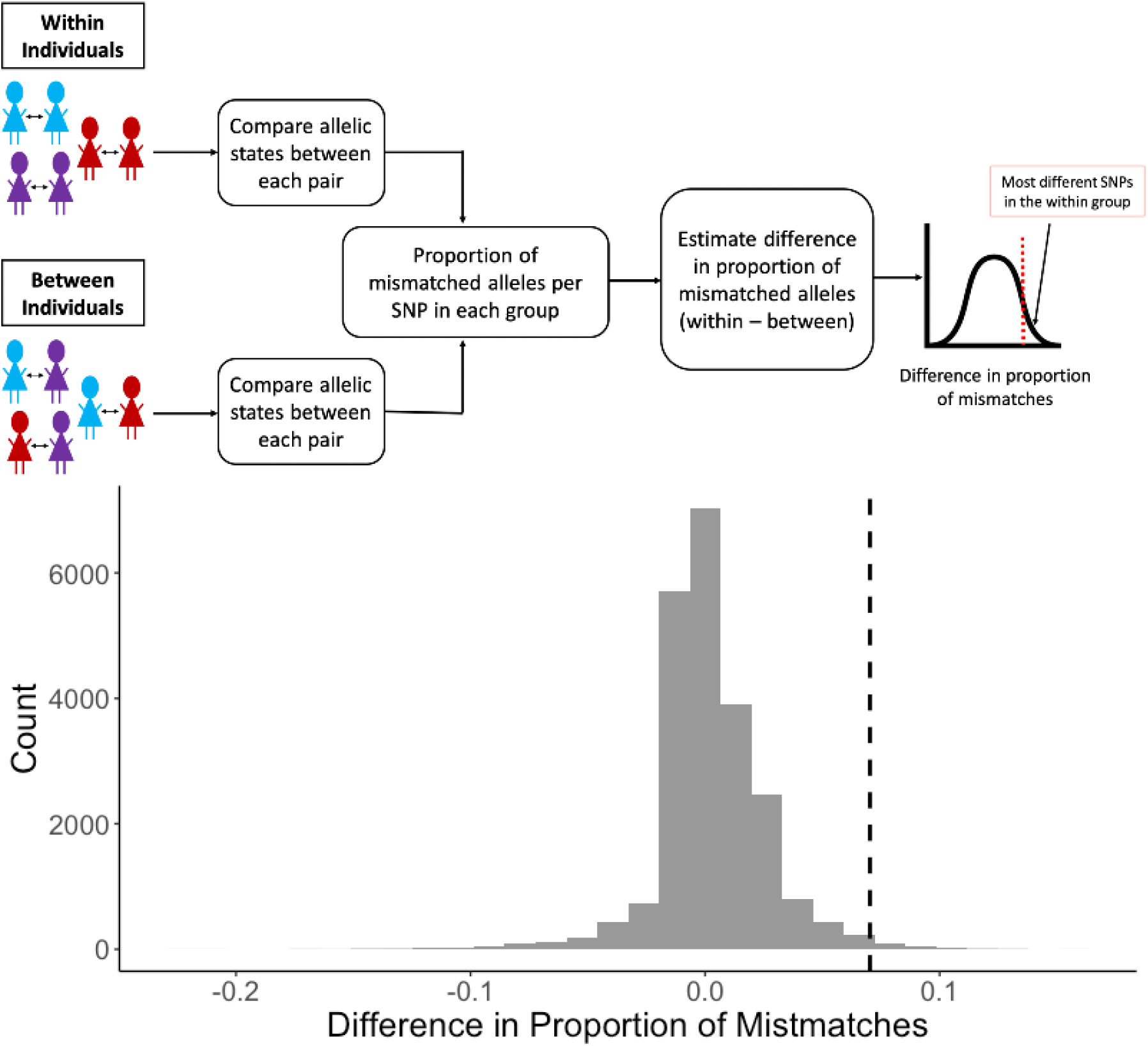
Analysis of mismatches in paired samples within and between individuals. a) Illustration of analysis to identify regions of the genome that vary more in parasites causing illness within the same individual over time (within individuals) compared to random pairs of parasites in the population (between individuals). b) Distribution of differences in the proportion of mismatched alleles in the within group and the between group. The difference was calculated as the proportion of mismatches at each non-synonymous SNP in the within group minus the proportion of mismatches at each non-synonymous SNP in the between group. The dashed black line indicates the threshold for the top 1% most different SNPs in the within group compared to the between group.

### Loci identified as likely targets of immunity by both analytical approaches

Twenty-five genes were identified by both analytical approaches, of which 11 (44%) encode proteins of unknown function (Table 2). Based on publicly available data, 20 of the 25 genes have a moderate to high level of expression in the erythrocytic stage of the parasite life cycle (Toenhake et al., 2018) (Supplementary Table 3). Eight genes have a mutagenesis index score near zero, suggesting that they are likely essential (Zhang et al., 2018), and at least 12 genes have either a known or a predicted transmembrane domain or a signal peptide (*PlasmoDB: A Functional Genomic Database for Malaria Parasites. - PubMed - NCBI*, n.d.). Among these 25 gene products are proteins whose putative function make them biologically plausible vaccine candidates, including SURFIN4.2 (thought to be involved in formation of the moving junction during erythrocyte invasion) (Quintana et al., 2018), as well as members of the CLAG and PHIST multigene families (both thought to have a role in parasite cytoadherence) (Holt et al., 1999; Proellocks et al., 2014).

**Table 2.**
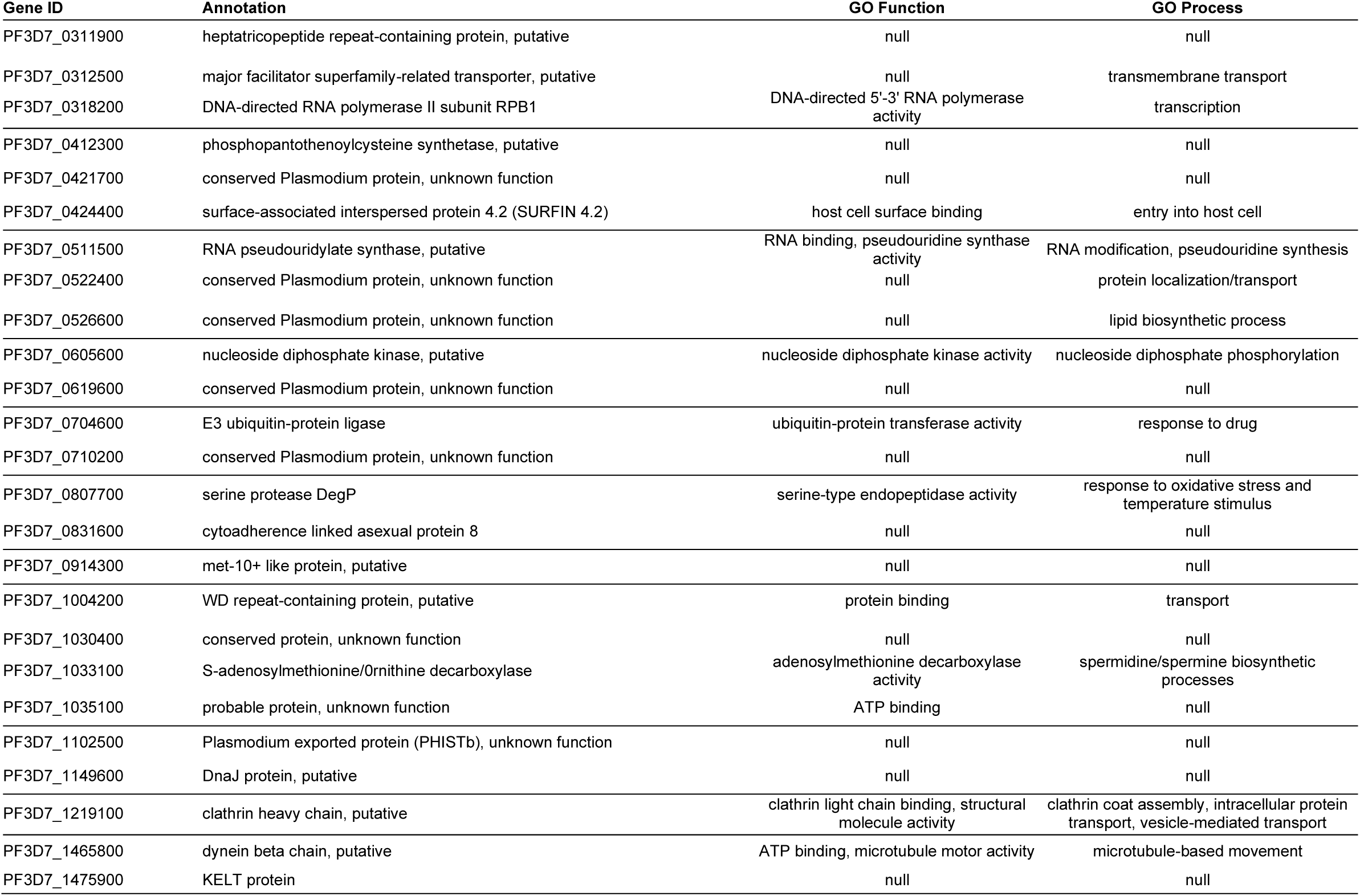
Gene products identified as likely targets of allele-specific immunity to malaria based on two analytical approaches.

As a proof of concept of the utility of our approach for identifying targets of allele-specific immunity, we used publicly available sequence data from Pf3K (*Pf3k Pilot Data Release 5* | *MalariaGEN*, n.d.) combined with 156 sequences from Papua New Guinea (PNG) from the MalariaGEN *P. falciparum* Community Project (MalariaGEN Plasmodium falciparum Community Project, 2016) to examine global diversity in *clag8*, one of the 25 identified genes that has been considered previously as a potential vaccine candidate, and found high nucleotide diversity, Tajima’s D, and number of segregating sites in the C-terminal region of *clag8*. This pattern was consistent when comparing data from parasites collected in Malawi to data from parasites from other regions of the world (Figure 4a). A haplotype network generated using *clag8* sequences from multiple geographic areas showed no evidence of regional adaptation (Figure 4b), including low *F*ST values between *clag8* sequences from Africa, Asia and PNG (Supplementary Table 5). Further analysis of CLAG8 protein sequences show regions with high protein disorder and B-cell epitope sites, especially in the C-terminal region (Supplementary Fig 4). To test our initial hypothesis that immune individuals become ill when infected with parasite antigen alleles that are rarer in the parasite population, we compared the frequency of *clag8* haplotypes based on the C-terminal region of the gene in individuals with different levels of immunity and observed that parasites causing illness in individuals in the high immunity group tended to have *clag8* haplotypes that were lower in frequency compared to individuals in the low immunity group, although significance was borderline (Supplementary Fig 5, *p*-value = 0.099, Wilcoxon rank sum test).

**Figure 4.**
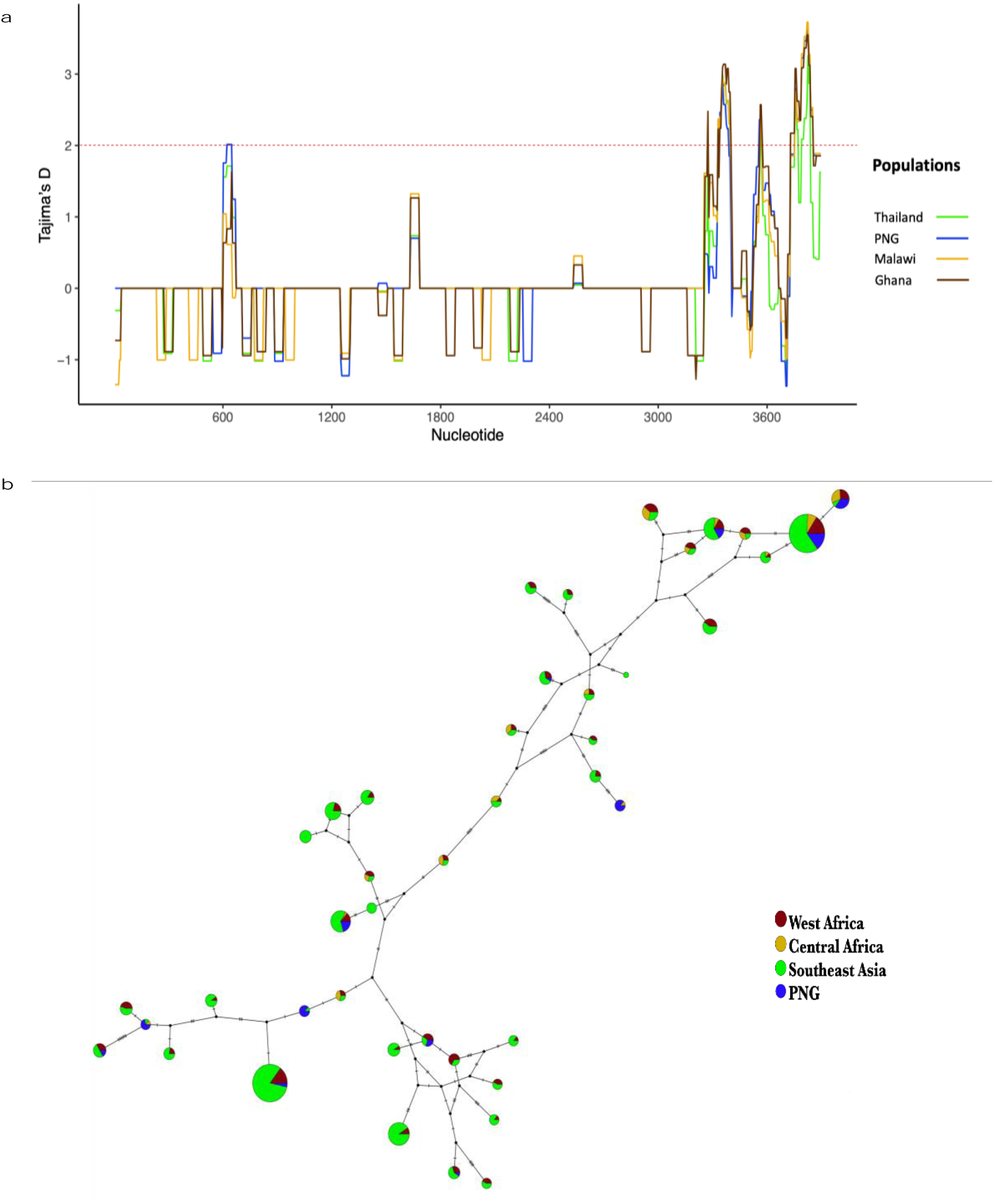
Global diversity of *clag8*. a) Tajima’s D along the *clag8* gene in samples from Thailand, PNG, Malawi, and Ghana. The dotted red lines represent a significant positive Tajima’s D value (≥ 2), suggestive of balancing selection. b) Haplotype network of *clag8* using *P. falciparum* sequence data.

## DISCUSSION

Previous studies have shown evidence of allele-specific acquisition of immunity to *P. falciparum* in single genes or proteins (Cortés et al., 2005; Early et al., 2018; Osier et al., 2008; Polley et al., 2007; Takala et al., 2007, 2009) that have been identified as potential vaccine antigens based on traditional vaccinology approaches that empirically identify immunogenic proteins. Other studies have performed genome-wide screens to identify genomic signatures of balancing selection but lack individual-level associations with clinical outcomes that might link specific signatures with protective immune responses (Amambua-Ngwa et al., 2012; Mobegi et al., 2014; Mu et al., 2007). In this study, we conducted a genome-wide, individual-level analysis to identify targets of allele-specific immunity to clinical malaria using *P. falciparum* whole genome sequencing data by identifying parasite genes that are genetically differentiated between individuals with different levels of immunity to malaria and genomic regions that are most different in parasites causing illness within the same individual versus between individuals. Twenty-five genes were identified using both analytical approaches and encode likely targets of allele-specific acquired immunity to clinical malaria, including genes thought to be involved in erythrocyte invasion and cytoadherence, among other functions. Examination of global diversity in *clag8*, a gene whose product has previously been considered as a vaccine candidate, provided evidence of immune selection in the C-terminal region of the gene and did not indicate geographical differences that might be indicative of local adaptation or genetic drift. Consistent with the hypothesis of allele-specific acquisition of immunity, *clag8* haplotypes of parasites causing illness in individuals with greater protective immunity were lower in frequency compared to those causing illness in less immune individuals. These findings support the utility of our approach for identification of targets of allele-specific immunity and the further investigation of these 25 loci as potential vaccine candidates.

In this study, we estimated the genetic complexity of infections and observed that individuals in the group with higher immunity generally had more complex infections than individuals with lower immunity. Although not statistically significant, this pattern is broadly consistent with results from other studies that have reported associations between infection complexity and either age and/or risk of disease (Färnert et al., 1999; Takala et al., 2007). The greater complexity of infections in individuals with higher immunity is consistent with the idea that immune individuals are capable of maintaining some parasite clones at a subclinical level and could suggest a possible role of polyclonal infections in maintaining immunity through continuous exposure to different strains (Bereczky et al., 2007; Sondén et al., 2015). In polyclonal infections from our data set, we observed that the predominant clone was more likely to have different alleles than the minor clone at sites identified as potential targets of immunity in our analysis. This finding is consistent with the hypothesis that minority clones are maintained at a subclinical level by acquired immunity, while the predominant clone is able to escape the immune response (because it has unrecognized alleles), resulting in a symptomatic infection. Prior to downstream analysis, infections lacking a predominant clone were excluded to avoid confounding by infection complexity and misclassification of clones likely responsible for disease symptoms. The exclusion of infections lacking a predominant clone led to the removal of a greater proportion of infections from the high immunity group than the low immunity group, which may have resulted in an underestimation of parasite diversity in the high immunity group. However, we do not believe this underestimation impacted our conclusions, as infections within the high immunity group were significantly more diverse at differentiated loci even after exclusion of these highly complex infections.

We hypothesized that individuals who have a higher degree of immunity to malaria would have a greater risk of malaria symptoms when infected with parasites having protein variants that are of lower frequency in the local parasite population, owing to their having already acquired immunity to variants encoded by more common alleles. When we estimated nucleotide diversity at differentiated loci in both immunity groups, we found significantly greater diversity in the group with higher immunity compared to the group with lower immunity. This difference in diversity may reflect the fact that individuals with higher immunity have symptomatic infections with parasite alleles that are rarer in the population and therefore more likely to be different from one another at differentiated loci. This scenario is also supported by our finding that the median frequency of the infecting allele was significantly lower in more immune individuals compared to less immune individuals when examining differentiated loci. These results also demonstrate the importance of accounting for rare alleles in vaccine design, as they may also lead to escape from vaccine-induced immunity.

Genome-wide screens for signatures of balancing selection have previously identified some of the 25 genes that we identified in this study, including PF3D7_0710200, *clag8* and other genes from the *surfin* and *phist* multigene families (Amambua-Ngwa et al., 2012; Mobegi et al., 2014; Mu et al., 2007). PF3D7_0710200, a gene encoding a conserved protein of unknown function, was identified by three separate studies as a potential immune target (Amambua-Ngwa et al., 2012; Mobegi et al., 2014; Mu et al., 2007); however, little is known about the function of this protein. Indeed, 11 of the 25 genes (44%) identified in this study encode for proteins of unknown function, highlighting the importance of further genetic screens to determine the role of such genes, which make up ∼35% of the parasite genome (Sexton et al., 2019). A recent study has suggested that most genes involved in host-parasite interactions, including many known antigens, are non-essential (Zhang et al., 2018); however, eight of the 25 genes identified in this study would be considered essential genes, based on their low mutagenesis index score (Zhang et al., 2018) (Supplementary Table 4). Such essential genes may be attractive vaccine targets as there may be less redundancy in function that would allow the parasite to adapt and escape vaccine-induced inhibition.

As a proof of concept of the utility of our approach for identifying targets of allele-specific immunity, we further examined the global diversity of one of the 25 identified genes, *clag8*, which has previously been considered as a malaria vaccine candidate. *clag8* belongs to the *clag* multigene family and is one of the least studied genes in the *clag* family. c*lag8* is highly expressed during the first few hours following erythrocyte invasion (early ring stage), displays decreased expression from 5 hours to 35 hours post invasion, and then increased expression during the schizont stage (Toenhake et al., 2018). It is thought to be part of the RhopH complex, formed by the members of *rhoph1/clag* gene families. The RhopH complex is an erythrocyte-binding protein complex inside the rhoptry and has been suggested to play an important role in establishment of the parasitophorous vacuole (Iriko et al., 2008; Kaneko et al., 2005; Sam-Yellowe & Perkins, 1991). Previous studies have reported evidence of positive diversifying selection in this gene, with high nucleotide diversity and a high proportion of non-synonymous substitutions per site (*d*N) (Iriko et al., 2008). Our global analysis of *clag8* diversity displayed high nucleotide diversity and Tajima’s D values in the C-terminal region of the gene, as well as evidence of high protein disorder and predicted B-cell epitopes in the C-terminal region of the protein, suggesting that it is likely to be immunogenic (Guy et al., 2015, p. 20). These results were consistent in all the countries included in the dataset, which represented parasite isolates from three major malaria endemic regions (*Pf3k Pilot Data Release 5* | *MalariaGEN*, n.d.). Additionally, *clag8* haplotypes displayed no evidence of geographical adaptation, with major haplotypes being observed in all geographic areas, and seemed to cluster into three main groups. Further studies to identify functional epitopes within the protein and potential cross-reactivity are necessary to determine whether related haplotypes can be grouped into serotypes for the purpose of designing a broadly protective vaccine. At an individual level, and in support of our overarching hypothesis, individuals with higher immunity to malaria were infected with *clag8* haplotypes that were less frequent compared to the *clag8* haplotypes infecting individuals with lower immunity.

It is noteworthy that our combined list of genes based on both analytical approaches did not include leading blood stage vaccine candidates such as AMA1 and the MSPs, although some of these genes were identified in a single approach. Using a protein microarray, Crompton *et al*. found that antibody responses to leading vaccine candidates, such as AMA1, MSP1, and MSP2, did not distinguish individuals who were protected from clinical infection versus those who were not in a cohort of individuals from Mali (Crompton et al., 2010), suggesting the possibility that responses to these proteins are not the primary drivers of clinical immunity. Other studies (Akpogheneta et al., 2008; Kinyanjui et al., 2007; Takala et al., 2009) have supported the hypothesis that responses to antigens such as AMA1 contribute to allele-specific clinical immunity but may be short-lived. In our analyses, infection pairs within an individual were not necessarily consecutive infections, with the time between infections ranging from 27 to 696 days. Although we did not see a significant correlation between the proportion of allele mismatches and the number of days between infections in a pair, it is possible that we could have failed to identify antigens involved in allele-specific acquisition of immunity that elicit short-lived immune responses. Studies with larger sample size would likely be required to distinguish antigens that have different antibody kinetics.

In addition, analyses in this study included only the core genome, owing to our use of selective whole genome amplification to enrich for parasite DNA prior to sequencing, which has been shown to result in poor sequencing coverage in the telomeric and centromeric regions (Oyola et al., 2016; Shah et al., 2020). Limiting analysis to the core genome could have prevented us from identifying members of multigene families that may be important for development of immunity to clinical malaria, as many of these genes are located in these low-coverage regions of the genome. However, because multigene families have been implicated in severe malaria, it is possible that immunity to these diverse antigens may be more relevant to preventing severe rather than uncomplicated malaria.

Here, we describe a promising genome-wide, individual-level approach to identify potential targets of allele-specific immunity to clinical malaria. Using this approach, we identified 25 genes, many of unknown function, that encode proteins that can be further characterized for their potential as candidates for a multivalent subunit malaria vaccine. Although further immunological validation will be necessary to confirm that the patterns observed in this study result from allele-specific immune responses, our results support the utility of this genomic epidemiology approach to identify new vaccine candidate antigens.

## METHODS

### Study design and samples

Parasite isolates were collected from participants in a longitudinal cohort study conducted in a rural area in southern Malawi. Details about the participants and study procedures have been described previously by Buchwald *et al* (Buchwald, Sixpence, et al., 2019). The data analyzed in this study were generated from red blood cell pellets collected from symptomatic, uncomplicated malaria infections. Samples were collected under protocols approved by the ethics committees at the College of Medicine in Blantyre, Malawi, and the University of Maryland, Baltimore, and with the informed consent of the participants or their guardians. The median parasitemia of the sampled infections as determined by microscopy was 21,960 parasites/μL and ranged from 0 parasites/μL (but positive by a rapid diagnostic test) to 241,260 parasites/μL. All samples were confirmed to be positive for *P. falciparum* via PCR. To ensure only independent infections were included in the analysis, infections within an individual separated by <14 days were excluded. DNA from red blood cell pellets was extracted using the method of Zainabadi *et al* (Zainabadi et al., 2017). Extracted DNA was enriched for parasite DNA using an optimized selective whole genome amplification approach described by Shah *et al* (Shah et al., 2020).

### Whole genome sequencing

Genomic DNA libraries were constructed for sequencing using the KAPA Library Preparation Kit (Kapa Biosystems, Woburn, MA). DNA (≥ 200 ηg) was fragmented with the Covaris E210 to ∼200 bp. Libraries were prepared using a modified version of the manufacturer’s protocol. The DNA was purified between enzymatic reactions and library size selection was performed with AMPure XT beads. Libraries were assessed for concentration and fragment size using the DNA High Sensitivity Assay on the LabChip GX (Perkin Elmer, Waltham, MA). Library concentrations were also assessed by qPCR using the KAPA Library Quantification Kit. Libraries were pooled and subsequently sequenced on an Illumina HiSeq 4000 (Illumina, San Diego, CA) to generate 150 bp paired-end reads. Sequencing data are available at NCBI under the project numbers and accession numbers listed in Supplementary Table 6.

### Read mapping and SNP Calling

Sequencing data were analyzed by mapping raw fastq files to the 3D7 reference genome using Bowtie2 (Langmead & Salzberg, 2012). Binary Alignment Map (BAM) files were processed following the GATK Best Practices workflow to obtain analysis-ready reads (DePristo et al., 2011; Van der Auwera et al., 2013). Bedtools (Quinlan & Hall, 2010) was used to generate coverage and depth estimates from the processed reads, and the GATK Best Practices workflow was followed for variant calling (DePristo et al., 2011; Van der Auwera et al., 2013). Haplotype Caller was used to create genomic variant call format (GVCF) files for each sample and joint SNP Calling was performed (GATK v3.7). Variants were removed if they met the following filtering criteria: variant confidence/quality by depth (QD) < 2.0, strand bias (FS) > 60.0, root mean square of the mapping quality (MQ) < 40.0, mapping quality rank sum (MQRankSum) < -12.5, read position rank sum (ReadPosRankSum) < -8.0, quality (QUAL) < 50. Variant sites with >20% missing genotypes and samples with >30% missing data were additionally removed using vcftools. Variants were also removed if the minor allele was not present in at least two samples. Only the core genome was used for further analysis. The median percentage of the genome covered with at least 20 reads was 88.87% (Shah et al., 2020).

### Definition of immune status

The degree of immunity to clinical malaria was defined based on the proportion of symptomatic infections out of all *P. falciparum* infections experienced by each study participant over the course of the two-year study. To account for exposure, individuals with less than five total infections, including symptomatic and asymptomatic infections, were excluded from the analysis. The median proportion of symptomatic infections was used as the cutoff to categorize individuals into higher and lower immunity groups.

### Complexity of infection and genetic differentiation

Only one infection from each individual was included in comparisons between high a low immunity groups. Infections were selected based on proximity to the median of the distribution of sampling dates to reduce temporal variability. DEploid-IBD (Zhu et al., 2017) was used to estimate the proportion of each clone within an infection. Infections without a predominant clone (i.e., where the majority clone had a frequency <60% within the infection) were defined as complex infections and were excluded from downstream analysis. For the remaining samples, the major allele was called at heterozygous positions if the allele was supported by ≥70% of reads; otherwise, the genotype was coded as missing. A Wilcoxon rank sum test was used to assess differences in the frequency of the majority clone in infections from the two immunity groups.

Vcftools (Danecek et al., 2011) was used to estimate Weir and Cockerham *F*ST in variable non-synonymous, bi-allelic sites. Significance was determined using 10,000 permutations, where the observed population was resampled without replacement. Nucleotide diversity at significantly differentiated sites was estimated using vcftools (Danecek et al., 2011). PlasmoDB (v44) (*PlasmoDB : The Plasmodium Genomics Resource*, n.d.) was used to identify genes containing differentiated SNPs.

In all polyclonal infections, the major and minor clones (defined by clone frequencies obtained from DEploid-IBD (Zhu et al., 2017)) were compared, provided clone frequency was less than 80% and greater than 10% (n = 23). At each non-synonymous site, the proportion of samples with mismatched alleles from major and minor clones was estimated. The proportion of mismatches was then compared between significantly differentiated sites and all remaining variable sites from the genome. The *p*-value was estimated by conducting a Wilcoxon rank sum test to determine if there is a significant difference in mismatches between clones at different sites *versus* remaining genome-wide variable sites.

### Paired infection analysis

Individuals with parasite whole genome sequence data from at least two symptomatic infections occurring at least 14 days apart were included in the comparison of infections occurring within the same host to infections occurring in different hosts. Multi-allelic sites were included in the analysis of paired infections, in contrast to analyses of genetic differentiation. The ‘within’ group included all pairs of parasites collected at different time points from the same individual. The ‘between group’, included all pairs of parasites from different individuals. A total of 116 samples were included in this study. The within group contained 124 pairs of samples and the between group contained 6546 pairs of samples. For all pairs, the allelic state was compared at each site and the proportion of pairs with non-matching allelic states was estimated by site (illustrated in Figure 3). The difference between the within group and the between group was calculated by subtracting the proportion of pairs with non-matching allelic states for each site. The *p-*value was estimated by conducting a one-sided z-test using the difference in proportion of mismatched alleles between the two groups. PlasmoDB (*PlasmoDB : The Plasmodium Genomics Resource*, n.d.) was used to identify genes containing the SNPs of interest.

### Global diversity in *clag8*

The MalariaGEN Pf3K project release 5.1 data(*Pf3k Pilot Data Release 5* | *MalariaGEN*, n.d.) was used to estimate global diversity in these genes identified in this study. The Pf3K dataset includes whole genome sequencing data from 2,512 samples collected in multiple locations in Asia and

Africa. Data from 156 additional isolates from Papua New Guinea were also included in the analysis. VaxPack (https://github.com/BarryLab01/vaxpack) was used for global population genetic analysis. GATKv4.0 was used for variant calling. Samples containing ambiguous bases were removed. Singleton SNPs were converted back to reference to prevent false positive variants. Nucleotide diversity and Tajima’s D were calculated for all polymorphic sites separately for every country that had a sample size greater than 50. Templeton, Crandall, and Sing (TCS) (Clement et al., 2002) method on PopArt (*Popart: Full-feature Software for Haplotype Network Construction - Leigh - 2015 - Methods in Ecology and Evolution - Wiley Online Library*, n.d.) was used to construct the haplotype network using non-synonymous SNPs. Protein disorder and B-cell epitopes were predicted using PlasmoSIP (Guy et al., 2015). The haplotype frequencies of the C-terminal region in Malawian isolates from different immunity groups were estimated for non-synonymous sites using DnaSP v6 (Rozas et al., 2017).

## Data Availability

Sequencing data are available at NCBI under the project numbers and accession numbers listed in Supplementary Table 6 of the manuscript.

## Acknowledgements

We thank the participants in the Mfera Cohort Study. We would like to thank Biraj Shrestha and Gillian Mbambo for their assistance with sample processing. We would also like to acknowledge Timothy D. O’Connor, Michael P. Cummings and Alexis Boleda for their valuable input and suggestions, and Terrie Taylor for her role in leading the Malawi International Center of Excellence for Malaria Research. This work was supported by funding from the following awards granted by the National Institutes of Health: R01AI101713, R01AI125579, R01AI141900, U19AI110820, K24AI114996, and the Malawi International Center of Excellence for Malaria Research U19AI089683.

## Author Contributions

Conceived and designed the study: ZS, MKL, JCS and ST-H. Collected and provided the samples: AGB, KBS, DPM, and MKL. Sequence processing and variant calling: ZS, KAM, and JCS. Analyzed the data: ZS, MTN, KAM, AD, DS, AEB, and ST-H. Interpreted the results: ZS, MA, AO, KBS, DS, AEB, MKL, JCS, and ST-H. Wrote the manuscript: ZS and ST-H. All authors have read and had the opportunity to comment on the manuscript.

## References

Akpogheneta, O. J., Duah, N. O., Tetteh, K. K. A., Dunyo, S., Lanar, D. E., Pinder, M., & Conway, D. J. (2008). Duration of Naturally Acquired Antibody Responses to Blood-Stage Plasmodium falciparum Is Age Dependent and Antigen Specific. Infection and Immunity, 76(4), 1748–1755. https://doi.org/10.1128/IAI.01333-07

Amambua-Ngwa, A., Tetteh, K. K. A., Manske, M., Gomez-Escobar, N., Stewart, L. B., Deerhake, M. E., Cheeseman, I. H., Newbold, C. I., Holder, A. A., Knuepfer, E., Janha, O., Jallow, M., Campino, S., MacInnis, B., Kwiatkowski, D. P., & Conway, D. J. (2012). Population Genomic Scan for Candidate Signatures of Balancing Selection to Guide Antigen Characterization in Malaria Parasites. PLoS Genetics, 8(11). https://doi.org/10.1371/journal.pgen.1002992

Barry, A. E., Trieu, A., Fowkes, F. J. I., Pablo, J., Kalantari-Dehaghi, M., Jasinskas, A., Tan, X., Kayala, M. A., Tavul, L., Siba, P. M., Day, K. P., Baldi, P., Felgner, P. L., & Doolan, D. L. (2011). The Stability and Complexity of Antibody Responses to the Major Surface Antigen of Plasmodium falciparum Are Associated with Age in a Malaria Endemic Area. Molecular & Cellular Proteomics, 10(11). https://doi.org/10.1074/mcp.M111.008326

Bejon, P., Williams, T. N., Liljander, A., Noor, A. M., Wambua, J., Ogada, E., Olotu, A., Osier, F. H. A., Hay, S. I., Färnert, A., & Marsh, K. (2010). Stable and unstable malaria hotspots in longitudinal cohort studies in Kenya. PLoS Medicine, 7(7), e1000304. https://doi.org/10.1371/journal.pmed.1000304

Bereczky, S., Liljander, A., Rooth, I., Faraja, L., Granath, F., Montgomery, S. M., & Färnert, A. (2007). Multiclonal asymptomatic Plasmodium falciparum infections predict a reduced risk of malaria disease in a Tanzanian population. Microbes and Infection, 9(1), 103– 110. https://doi.org/10.1016/j.micinf.2006.10.014

Bhatia, G., Patterson, N., Sankararaman, S., & Price, A. L. (2013). Estimating and interpreting FST: The impact of rare variants. Genome Research, 23(9), 1514–1521. https://doi.org/10.1101/gr.154831.113

Bousema, T., Drakeley, C., Gesase, S., Hashim, R., Magesa, S., Mosha, F., Otieno, S., Carneiro, I., Cox, J., Msuya, E., Kleinschmidt, I., Maxwell, C., Greenwood, B., Riley, E., Sauerwein, R., Chandramohan, D., & Gosling, R. (2010). Identification of Hot Spots of Malaria Transmission for Targeted Malaria Control. The Journal of Infectious Diseases, 201(11), 1764–1774. https://doi.org/10.1086/652456

Buchwald, A. G., Sixpence, A., Chimenya, M., Damson, M., Sorkin, J. D., Wilson, M. L., Seydel, K., Hochman, S., Mathanga, D. P., Taylor, T. E., & Laufer, M. K. (2019). Clinical Implications of Asymptomatic Plasmodium falciparum Infections in Malawi. Clinical Infectious Diseases: An Official Publication of the Infectious Diseases Society of America, 68(1), 106–112. https://doi.org/10.1093/cid/ciy427

Buchwald, A. G., Sorkin, J. D., Sixpence, A., Chimenya, M., Damson, M., Wilson, M. L., Seydel, K., Hochman, S., Mathanga, D., Taylor, T. E., & Laufer, M. K. (2019). Association Between Age and Plasmodium falciparum Infection Dynamics. American Journal of Epidemiology, 188(1), 169–176. https://doi.org/10.1093/aje/kwy213

Clark, T. D., Greenhouse, B., Njama-Meya, D., Nzarubara, B., Maiteki-Sebuguzi, C., Staedke, S. G., Seto, E., Kamya, M. R., Rosenthal, P. J., & Dorsey, G. (2008). Factors determining the heterogeneity of malaria incidence in children in Kampala, Uganda. The Journal of Infectious Diseases, 198(3), 393–400. https://doi.org/10.1086/589778

Clement, M., Snell, Q., Walke, P., Posada, D., & Crandall, K. (2002). TCS: Estimating gene genealogies. Proceedings 16th International Parallel and Distributed Processing Symposium, 7 pp. https://doi.org/10.1109/IPDPS.2002.1016585

Cortés, A., Mellombo, M., Masciantonio, R., Murphy, V. J., Reeder, J. C., & Anders, R. F. (2005). Allele specificity of naturally acquired antibody responses against Plasmodium falciparum apical membrane antigen 1. Infection and Immunity, 73(1), 422–430. https://doi.org/10.1128/IAI.73.1.422-430.2005

Cowman, A. F., Healer, J., Marapana, D., & Marsh, K. (2016). Malaria: Biology and Disease. Cell, 167(3), 610–624. https://doi.org/10.1016/j.cell.2016.07.055

Crompton, P. D., Kayala, M. A., Traore, B., Kayentao, K., Ongoiba, A., Weiss, G. E., Molina, D. M., Burk, C. R., Waisberg, M., Jasinskas, A., Tan, X., Doumbo, S., Doumtabe, D., Kone, Y., Narum, D. L., Liang, X., Doumbo, O. K., Miller, L. H., Doolan, D. L., … Pierce, S. K. (2010). A prospective analysis of the Ab response to Plasmodium falciparum before and after a malaria season by protein microarray. Proceedings of the National Academy of Sciences, 107(15), 6958–6963. https://doi.org/10.1073/pnas.1001323107

Danecek, P., Auton, A., Abecasis, G., Albers, C. A., Banks, E., DePristo, M. A., Handsaker, R. E., Lunter, G., Marth, G. T., Sherry, S. T., McVean, G., Durbin, R., & 1000 Genomes Project Analysis Group. (2011). The variant call format and VCFtools. Bioinformatics (Oxford, England), 27(15), 2156–2158. https://doi.org/10.1093/bioinformatics/btr330

DePristo, M. A., Banks, E., Poplin, R., Garimella, K. V., Maguire, J. R., Hartl, C., Philippakis, A. A., del Angel, G., Rivas, M. A., Hanna, M., McKenna, A., Fennell, T. J., Kernytsky, A. M., Sivachenko, A. Y., Cibulskis, K., Gabriel, S. B., Altshuler, D., & Daly, M. J. (2011). A framework for variation discovery and genotyping using next-generation DNA sequencing data. Nature Genetics, 43(5), 491–498. https://doi.org/10.1038/ng.806

Dutta, S., Lee, S. Y., Batchelor, A. H., & Lanar, D. E. (2007). Structural basis of antigenic escape of a malaria vaccine candidate. Proceedings of the National Academy of Sciences, 104(30), 12488–12493. https://doi.org/10.1073/pnas.0701464104

Early, A. M., Lievens, M., MacInnis, B. L., Ockenhouse, C. F., Volkman, S. K., Adjei, S., Agbenyega, T., Ansong, D., Gondi, S., Greenwood, B., Hamel, M., Odero, C., Otieno, K., Otieno, W., Owusu-Agyei, S., Asante, K. P., Sorgho, H., Tina, L., Tinto, H., … Neafsey, D. E. (2018). Host-mediated selection impacts the diversity of Plasmodium falciparum antigens within infections. Nature Communications, 9. https://doi.org/10.1038/s41467-018-03807-7

Egan, A. F., Morris, J., Barnish, G., Allen, S., Greenwood, B. M., Kaslow, D. C., Holder, A. A., & Riley, E. M. (1996). Clinical immunity to Plasmodium falciparum malaria is associated with serum antibodies to the 19-kDa C-terminal fragment of the merozoite surface antigen, PfMSP-1. The Journal of Infectious Diseases, 173(3), 765–769. https://doi.org/10.1093/infdis/173.3.765

Elissa, N., Migot-Nabias, F., Luty, A., Renaut, A., Touré, F., Vaillant, M., Lawoko, M., Yangari, P., Mayombo, J., Lekoulou, F., Tshipamba, P., Moukagni, R., Millet, P., & Deloron, P. (2003). Relationship between entomological inoculation rate, Plasmodium falciparum prevalence rate, and incidence of malaria attack in rural Gabon. Acta Tropica, 85(3), 355–361. https://doi.org/10.1016/s0001-706x(02)00266-8

Färnert, A., Rooth, I., Svensson, Å., Snounou, G., & Björkman, A. (1999). Complexity of Plasmodium falciparum Infections Is Consistent over Time and Protects against Clinical Disease in Tanzanian Children. The Journal of Infectious Diseases, 179(4), 989–995. https://doi.org/10.1086/314652

Gardner, M. J., Hall, N., Fung, E., White, O., Berriman, M., Hyman, R. W., Carlton, J. M., Pain, A., Nelson, K. E., Bowman, S., Paulsen, I. T., James, K., Eisen, J. A., Rutherford, K., Salzberg, S. L., Craig, A., Kyes, S., Chan, M.-S., Nene, V., … Barrell, B. (2002). Genome sequence of the human malaria parasite Plasmodium falciparum. Nature, 419(6906), 498–511. https://doi.org/10.1038/nature01097

Gaudart, J., Poudiougou, B., Dicko, A., Ranque, S., Toure, O., Sagara, I., Diallo, M., Diawara, S., Ouattara, A., Diakite, M., & Doumbo, O. K. (2006). Space-time clustering of childhood malaria at the household level: A dynamic cohort in a Mali village. BMC Public Health, 6, 286. https://doi.org/10.1186/1471-2458-6-286

Gupta, A., Thiruvengadam, G., & Desai, S. A. (2015). The conserved clag multigene family of malaria parasites: Essential roles in host-pathogen interaction. Drug Resistance Updates?: Reviews and Commentaries in Antimicrobial and Anticancer Chemotherapy, 18, 47–54. https://doi.org/10.1016/j.drup.2014.10.004

Guy, A. J., Irani, V., MacRaild, C. A., Anders, R. F., Norton, R. S., Beeson, J. G., Richards, J. S., & Ramsland, P. A. (2015). Insights into the Immunological Properties of Intrinsically Disordered Malaria Proteins Using Proteome Scale Predictions. PLOS ONE, 10(10), e0141729. https://doi.org/10.1371/journal.pone.0141729

Holt, D. C., Gardiner, D. L., Thomas, E. A., Mayo, M., Bourke, P. F., Sutherland, C. J., Carter, R., Myers, G., Kemp, D. J., & Trenholme, K. R. (1999). The cytoadherence linked asexual gene family of Plasmodium falciparum: Are there roles other than cytoadherence? International Journal for Parasitology, 29(6), 939–944. https://doi.org/10.1016/s0020-7519(99)00046-6

Iriko, H., Kaneko, O., Otsuki, H., Tsuboi, T., Su, X., Tanabe, K., & Torii, M. (2008). Diversity and evolution of the rhoph1/clag multigene family of Plasmodium falciparum. Molecular and Biochemical Parasitology, 158(1), 11–21. https://doi.org/10.1016/j.molbiopara.2007.11.004

Kaneko, O., Lim, B. Y. S. Y., Iriko, H., Ling, I. T., Otsuki, H., Grainger, M., Tsuboi, T., Adams, J. H., Mattei, D., Holder, A. A., & Torii, M. (2005). Apical expression of three RhopH1/Clag proteins as components of the Plasmodium falciparum RhopH complex. Molecular and Biochemical Parasitology, 143(1), 20–28. https://doi.org/10.1016/j.molbiopara.2005.05.003

Kang, S. Y., Battle, K. E., Gibson, H. S., Cooper, L. V., Maxwell, K., Kamya, M., Lindsay, S. W., Dorsey, G., Greenhouse, B., Rodriguez-Barraquer, I., Reiner, R. C. Jr., Smith, D. L., & Bisanzio, D. (2018). Heterogeneous exposure and hotspots for malaria vectors at three study sites in Uganda. Gates Open Research, 2. https://doi.org/10.12688/gatesopenres.12838.2

Kinyanjui, S. M., Conway, D. J., Lanar, D. E., & Marsh, K. (2007). IgG antibody responses to Plasmodium falciparum merozoite antigens in Kenyan children have a short half-life. Malaria Journal, 6, 82. https://doi.org/10.1186/1475-2875-6-82

Krzyczmonik, K., Świtnicki, M., & Kaczanowski, S. (2012). Analysis of immunogenicity of different protein groups from malaria parasite Plasmodium falciparum. Infection, Genetics and Evolution, 12(8), 1911–1916. https://doi.org/10.1016/j.meegid.2012.07.023

Langmead, B., & Salzberg, S. L. (2012). Fast gapped-read alignment with Bowtie 2. Nature Methods, 9(4), 357–359. https://doi.org/10.1038/nmeth.1923

MalariaGEN Plasmodium falciparum Community Project. (2016). Genomic epidemiology of artemisinin resistant malaria. ELife, 5, e08714. https://doi.org/10.7554/eLife.08714

Marsh, K., & Kinyanjui, S. (2006). Immune effector mechanisms in malaria. Parasite Immunology, 28(1–2), 51–60. https://doi.org/10.1111/j.1365-3024.2006.00808.x

Metzger, W. G., Okenu, D. M. N., Cavanagh, D. R., Robinson, J. V., Bojang, K. A., Weiss, H. A., McBride, J. S., Greenwood, B. M., & Conway, D. J. (2003). Serum IgG3 to the Plasmodium falciparum merozoite surface protein 2 is strongly associated with a reduced prospective risk of malaria. Parasite Immunology, 25(6), 307–312.

Mobegi, V. A., Duffy, C. W., Amambua-Ngwa, A., Loua, K. M., Laman, E., Nwakanma, D. C., MacInnis, B., Aspeling-Jones, H., Murray, L., Clark, T. G., Kwiatkowski, D. P., & Conway, D. J. (2014). Genome-wide analysis of selection on the malaria parasite Plasmodium falciparum in West African populations of differing infection endemicity. Molecular Biology and Evolution, 31(6), 1490–1499. https://doi.org/10.1093/molbev/msu106

Mu, J., Awadalla, P., Duan, J., McGee, K. M., Keebler, J., Seydel, K., McVean, G. A. T., & Su, X. (2007). Genome-wide variation and identification of vaccine targets in the Plasmodium falciparum genome. Nature Genetics, 39(1), 126–130. https://doi.org/10.1038/ng1924

Osier, F. H. A., Fegan, G., Polley, S. D., Murungi, L., Verra, F., Tetteh, K. K. A., Lowe, B., Mwangi, T., Bull, P. C., Thomas, A. W., Cavanagh, D. R., McBride, J. S., Lanar, D. E., Mackinnon, M. J., Conway, D. J., & Marsh, K. (2008). Breadth and magnitude of antibody responses to multiple Plasmodium falciparum merozoite antigens are associated with protection from clinical malaria. Infection and Immunity, 76(5), 2240– 2248. https://doi.org/10.1128/IAI.01585-07

Ouattara, A., Barry, A. E., Dutta, S., Remarque, E. J., Beeson, J. G., & Plowe, C. V. (2015). Designing malaria vaccines to circumvent antigen variability. Vaccine, 33(52), 7506– 7512. https://doi.org/10.1016/j.vaccine.2015.09.110

Ouattara, A., Takala-Harrison, S., Thera, M. A., Coulibaly, D., Niangaly, A., Saye, R., Tolo, Y., Dutta, S., Heppner, D. G., Soisson, L., Diggs, C. L., Vekemans, J., Cohen, J., Blackwelder, W. C., Dube, T., Laurens, M. B., Doumbo, O. K., & Plowe, C. V. (2013). Molecular basis of allele-specific efficacy of a blood-stage malaria vaccine: Vaccine development implications. The Journal of Infectious Diseases, 207(3), 511–519. https://doi.org/10.1093/infdis/jis709

Oyola, S. O., Ariani, C. V., Hamilton, W. L., Kekre, M., Amenga-Etego, L. N., Ghansah, A., Rutledge, G. G., Redmond, S., Manske, M., Jyothi, D., Jacob, C. G., Otto, T. D., Rockett, K., Newbold, C. I., Berriman, M., & Kwiatkowski, D. P. (2016). Whole genome sequencing of Plasmodium falciparum from dried blood spots using selective whole genome amplification. Malaria Journal, 15. https://doi.org/10.1186/s12936-016-1641-7

Perraut, R., Varela, M.-L., Joos, C., Diouf, B., Sokhna, C., Mbengue, B., Tall, A., Loucoubar, C., Touré, A., & Mercereau-Puijalon, O. (2017). Association of antibodies to Plasmodium falciparum merozoite surface protein-4 with protection against clinical malaria. Vaccine, 35(48 Pt B), 6720–6726. https://doi.org/10.1016/j.vaccine.2017.10.012

Pf3k pilot data release 5 | MalariaGEN. (n.d.). Retrieved December 23, 2019, from https://www.malariagen.net/data/pf3K-5

PlasmoDB: a functional genomic database for malaria parasites. -PubMed—NCBI. (n.d.). Retrieved February 18, 2020, from https://www.ncbi.nlm.nih.gov/pubmed?cmd=search&term=18957442

PlasmoDB?: The Plasmodium Genomics Resource. (n.d.). Retrieved May 1, 2018, from http://plasmodb.org/plasmo/

Polley, S. D., Tetteh, K. K. A., Lloyd, J. M., Akpogheneta, O. J., Greenwood, B. M., Bojang, K. A., & Conway, D. J. (2007). Plasmodium falciparum Merozoite Surface Protein 3 Is a Target of Allele-Specific Immunity and Alleles Are Maintained by Natural Selection. The Journal of Infectious Diseases, 195(2), 279–287. https://doi.org/10.1086/509806

Popart: Full-feature software for haplotype network construction—Leigh—2015—Methods in Ecology and Evolution—Wiley Online Library. (n.d.). Retrieved February 4, 2020, from https://besjournals.onlinelibrary.wiley.com/doi/full/10.1111/2041-210X.12410

Portugal, S., Pierce, S. K., & Crompton, P. D. (2013). Young Lives Lost as B Cells Falter: What We Are Learning About Antibody Responses in Malaria. The Journal of Immunology, 190(7), 3039–3046. https://doi.org/10.4049/jimmunol.1203067

Proellocks, N. I., Herrmann, S., Buckingham, D. W., Hanssen, E., Hodges, E. K., Elsworth, B., Morahan, B. J., Coppel, R. L., & Cooke, B. M. (2014). A lysine-rich membrane-associated PHISTb protein involved in alteration of the cytoadhesive properties of Plasmodium falciparum-infected red blood cells. FASEB Journal: Official Publication of the Federation of American Societies for Experimental Biology, 28(7), 3103–3113. https://doi.org/10.1096/fj.14-250399

Quinlan, A. R., & Hall, I. M. (2010). BEDTools: A flexible suite of utilities for comparing genomic features. Bioinformatics, 26(6), 841–842. https://doi.org/10.1093/bioinformatics/btq033

Quintana, M. D. P., Ch’ng, J.-H., Zandian, A., Imam, M., Hultenby, K., Theisen, M., Nilsson, P., Qundos, U., Moll, K., Chan, S., & Wahlgren, M. (2018). SURGE complex of Plasmodium falciparum in the rhoptry-neck (SURFIN4.2-RON4-GLURP) contributes to merozoite invasion. PloS One, 13(8), e0201669. https://doi.org/10.1371/journal.pone.0201669

Rozas, J., Ferrer-Mata, A., Sánchez-DelBarrio, J. C., Guirao-Rico, S., Librado, P., Ramos-Onsins, S. E., & Sánchez-Gracia, A. (2017). DnaSP 6: DNA Sequence Polymorphism Analysis of Large Data Sets. Molecular Biology and Evolution, 34(12), 3299–3302. https://doi.org/10.1093/molbev/msx248

Ryg-Cornejo, V., Ly, A., & Hansen, D. S. (2016). Immunological processes underlying the slow acquisition of humoral immunity to malaria. Parasitology, 143(2), 199–207. https://doi.org/10.1017/S0031182015001705

Sam-Yellowe, T. Y., & Perkins, M. E. (1991). Interaction of the 140/130/110 kDa rhoptry protein complex of Plasmodium falciparum with the erythrocyte membrane and liposomes. Experimental Parasitology, 73(2), 161–171. https://doi.org/10.1016/0014-4894(91)90019-S

Sexton, A. E., Doerig, C., Creek, D. J., & Carvalho, T. G. (2019). Post-Genomic Approaches to Understanding Malaria Parasite Biology: Linking Genes to Biological Functions. ACS Infectious Diseases, 5(8), 1269–1278. https://doi.org/10.1021/acsinfecdis.9b00093

Shah, Z., Adams, M., Moser, K. A., Shrestha, B., Stucke, E. M., Laufer, M. K., Serre, D., Silva, J. C., & Takala-Harrison, S. (2020). Optimization of parasite DNA enrichment approaches to generate whole genome sequencing data for Plasmodium falciparum from low parasitaemia samples. Malaria Journal, 19(1), 135. https://doi.org/10.1186/s12936-020-03195-8

Sondén, K., Doumbo, S., Hammar, U., Vafa Homann, M., Ongoiba, A., Traoré, B., Bottai, M., Crompton, P. D., & Färnert, A. (2015). Asymptomatic Multiclonal Plasmodium falciparum Infections Carried Through the Dry Season Predict Protection Against Subsequent Clinical Malaria. The Journal of Infectious Diseases, 212(4), 608–616. https://doi.org/10.1093/infdis/jiv088

Takala, S. L., Coulibaly, D., Thera, M. A., Batchelor, A. H., Cummings, M. P., Escalante, A. A., Ouattara, A., Traoré, K., Niangaly, A., Djimdé, A. A., Doumbo, O. K., & Plowe, C. V. (2009). Extreme Polymorphism in a Vaccine Antigen and Risk of Clinical Malaria: Implications for Vaccine Development. Science Translational Medicine, 1(2), 2ra5–2ra5. https://doi.org/10.1126/scitranslmed.3000257

Takala, S. L., Coulibaly, D., Thera, M. A., Dicko, A., Smith, D. L., Guindo, A. B., Kone, A. K., Traore, K., Ouattara, A., Djimde, A. A., Sehdev, P. S., Lyke, K. E., Diallo, D. A., Doumbo, O. K., & Plowe, C. V. (2007). Dynamics of polymorphism in a malaria vaccine antigen at a vaccine-testing site in Mali. PLoS Medicine, 4(3), e93. https://doi.org/10.1371/journal.pmed.0040093

Thera, M. A., Doumbo, O. K., Coulibaly, D., Laurens, M. B., Ouattara, A., Kone, A. K., Guindo, A. B., Traore, K., Traore, I., Kouriba, B., Diallo, D. A., Diarra, I., Daou, M., Dolo, A., Tolo, Y., Sissoko, M. S., Niangaly, A., Sissoko, M., Takala-Harrison, S., … Plowe, C. V. (2011). A field trial to assess a blood-stage malaria vaccine. The New England Journal of Medicine, 365(11), 1004–1013. https://doi.org/10.1056/NEJMoa1008115

Toenhake, C. G., Fraschka, S. A.-K., Vijayabaskar, M. S., Westhead, D. R., van Heeringen, S. J., & Bártfai, R. (2018). Chromatin Accessibility-Based Characterization of the Gene Regulatory Network Underlying Plasmodium falciparum Blood-Stage Development. Cell Host & Microbe, 23(4), 557-569.e9. https://doi.org/10.1016/j.chom.2018.03.007

Tran, T. M., Ongoiba, A., Coursen, J., Crosnier, C., Diouf, A., Huang, C.-Y., Li, S., Doumbo, S., Doumtabe, D., Kone, Y., Bathily, A., Dia, S., Niangaly, M., Dara, C., Sangala, J., Miller, L. H., Doumbo, O. K., Kayentao, K., Long, C. A., … Crompton, P. D. (2014). Naturally Acquired Antibodies Specific for Plasmodium falciparum Reticulocyte-Binding Protein Homologue 5 Inhibit Parasite Growth and Predict Protection From Malaria. The Journal of Infectious Diseases, 209(5), 789–798. https://doi.org/10.1093/infdis/jit553

Van der Auwera, G. A., Carneiro, M. O., Hartl, C., Poplin, R., Del Angel, G., Levy-Moonshine, A., Jordan, T., Shakir, K., Roazen, D., Thibault, J., Banks, E., Garimella, K. V., Altshuler, D., Gabriel, S., & DePristo, M. A. (2013). From FastQ data to high confidence variant calls: The Genome Analysis Toolkit best practices pipeline. Current Protocols in Bioinformatics, 43, 11.10.1-33. https://doi.org/10.1002/0471250953.bi1110s43

van Schaijk, B. C. L., Ploemen, I. H. J., Annoura, T., Vos, M. W., Foquet, L., van Gemert, G.-J., Chevalley-Maurel, S., van de Vegte-Bolmer, M., Sajid, M., Franetich, J.-F., Lorthiois, A., Leroux-Roels, G., Meuleman, P., Hermsen, C. C., Mazier, D., Hoffman, S. L., Janse, C. J., Khan, S. M., & Sauerwein, R. W. (2014). A genetically attenuated malaria vaccine candidate based on P. falciparum b9/slarp gene-deficient sporozoites. ELife, 3. https://doi.org/10.7554/eLife.03582

Vanegas, M., Bermúdez, A., Guerrero, Y. A., Cortes-Vecino, J. A., Curtidor, H., Patarroyo, M. E., & Lozano, J. M. (2014). Protecting capacity against malaria of chemically defined tetramer forms based on the Plasmodium falciparum apical sushi protein as potential vaccine components. Biochemical and Biophysical Research Communications, 451(1), 15–23. https://doi.org/10.1016/j.bbrc.2014.06.143

Weedall, G. D., & Conway, D. J. (2010). Detecting signatures of balancing selection to identify targets of anti-parasite immunity. Trends in Parasitology, 26(7), 363–369. https://doi.org/10.1016/j.pt.2010.04.002

WHO | World malaria report 2018. (n.d.). WHO. Retrieved May 30, 2019, from http://www.who.int/malaria/publications/world-malaria-report-2018/report/en/

Zainabadi, K., Adams, M., Han, Z. Y., Lwin, H. W., Han, K. T., Ouattara, A., Thura, S., Plowe, C. V., & Nyunt, M. M. (2017). A novel method for extracting nucleic acids from dried blood spots for ultrasensitive detection of low-density Plasmodium falciparum and Plasmodium vivax infections. Malaria Journal, 16(1), 377. https://doi.org/10.1186/s12936-017-2025-3

Zhang, M., Wang, C., Otto, T. D., Oberstaller, J., Liao, X., Adapa, S. R., Udenze, K., Bronner, I. F., Casandra, D., Mayho, M., Brown, J., Li, S., Swanson, J., Rayner, J. C., Jiang, R. H. Y., & Adams, J. H. (2018). Uncovering the essential genes of the human malaria parasite Plasmodium falciparum by saturation mutagenesis. Science, 360(6388). https://doi.org/10.1126/science.aap7847

Zhu, S. J., Almagro-Garcia, J., & McVean, G. (2017). Deconvolution of multiple infections in Plasmodium falciparum from high throughput sequencing data. Bioinformatics (Oxford, England). https://doi.org/10.1093/bioinformatics/btx530

